# Reproductive Maternal and Newborn Health providers assessment of facility preparedness and its Determinants during the COVID-19 pandemic in Lagos, Nigeria

**DOI:** 10.1101/2020.09.24.20201319

**Authors:** Ameh Charles, Banke-Thomas Aduragbemi, Balogun Mobolanle, Makwe Christian Chigozie, Afolabi Bosede Bukola

## Abstract

The global COVID-19 pandemic is predicted to compromise the achievement of global reproductive, maternal and newborn health (RMNH) targets. The objective of this study was to determine the health facility (HF) preparedness for RMNH service delivery during the outbreak from the perspective of RMNH providers and to determine what factors significantly predict this. An anonymous cross-sectional online survey of RMNH providers was conducted from 1^st^ to 21^st^ July 2020 in Lagos state Nigeria. We conducted a descriptive and ordinal regression analysis, with RMNH worker perception of HF preparedness for RMNH service delivery during the outbreak as the dependent variable.

Two hundred and fifty-six RMNH workers participated, 35.2% reported that RMNH services were unavailable at some time since March 2020, 39% felt moderate or extreme work-related burnout, 84% were moderately or extremely concerned about the availability of PPE and related guidelines, and only 11.7% were extremely satisfied with the preparedness of their HFs. Our final model was a statistically significant predictor of RMNH worker perception of HF preparedness explaining 54.7% of the variation in the outcome variable. A one-unit increase in the level of satisfaction with the communication from HF management and level of concern about the availability of PPE and COVID-19 guidelines would increase the odds of observing a higher category of satisfaction with HF COVID-19 preparedness (OR 0.79-2.92, p<0.001 and 0.02-0.15 p<0.001 respectively).

Adequate support of RMNH providers particularly provision of PPE and guidelines, appropriate communications about COVID- 19 should be prioritised as part of health system preparedness.

## Introduction

The World Health Organization declared Coronavirus disease (COVID-19) as a pandemic on 11^th^ March 2020, after first being reported in Hubei Province, China in December 2019. As of 20^th^ September 2020, over 30 million cases and close to 1 million deaths have been recorded globally, with Brazil, India and the United States of America accounting for about 50% of the cases.^1^ For the same period, 1.2 million cases and over 30, 000 deaths were reported from Africa, with the highest numbers from South Africa, Egypt, Morocco, Ethiopia and Nigeria.^2^

Governments around the world have implemented various public health and social measures to reduce the spread of COVID-19. Health service provision has been modified in many settings to focus on managing COVID-19 cases, by reducing service provision for non-COVID-19 health emergencies and essential health services. Thus, health systems have struggled to cope with maintaining essential health services while trying to control the infection.^3^ Of all health services, RMNH services, are likely to be impacted the most, like in previous infectious disease pandemic. A systematic review of the impact of the 2014-15 Ebola outbreak reported that the health system in affected areas collapsed due to overwhelming demand directly linked to the outbreak, health workers death, resource diversion and closure of facilities that comprised access to essential health services. They also reported that in Ebola-affected areas, there was an 80% reduction in maternal delivery care and increased morbidity and mortality.^4^

A recent study that modelled the impact of workforce, supplies, demand and access reductions due to the COVID-19 outbreak showed that 9.8%-51.9% reduction in coverage could result in up to 113, 000 additional maternal deaths (38.6% increase from baseline) in 12 months, in 118 low and middle-income countries. ^5^ Sixty per cent of these estimated deaths may be due to a reduction in the availability of drugs to prevent and treat bleeding after birth, treat infections, treat pregnancy-related convulsions and lack of clean birth environments. ^5^ Also, Riley, et al. estimated the potential impact of COVID-19 pandemic on sexual and reproductive health in low-and-middle-income countries, reporting that a 10% decline in the use of short and long-acting contraceptives will result in 15.4 million unintended pregnancies.^6^ While the mechanism for the reduction in supplies, demand and access can be linked to the direct effect of various policies that restrict movement resulting in a disruption in the supply chain, household income and travel infrastructure, the mechanism for its influence on the health workforce is not straightforward. There are several possibilities including but not limited to staff COVID-19 infection/and death, redeployment to COVID-19 treatment units, staff discrimination of infected patients, staff refusal to treat COVID-19 patients, staff experiencing community discrimination, work-related stress resulting in absenteeism or staff off work due to a need to care for infected family members.^3,7^

Before the pandemic, Nigeria already contributed to about 13% of the estimated global maternal deaths annually and had an estimated maternal mortality ratio (MMR) of 556/100, 000 live births, achieving the target of 140/100,000 live births as part of the ending preventable maternal mortality over the next 10 years appears unrealistic.^8^ Now within the context of the COVID-19 pandemic, the sufficiency of RMNH services in a state like Lagos which is the epicentre of the disease comes into sharp focus. According to the Nigeria Centres for Disease Control, as of September 20^th^, 2020, there are over 57,000 confirmed cases and 1,100 deaths with Lagos State accounting for about a 33% of all cases (19,055) and 18% (205) of all deaths. ^9^

With the health workers being the ones at the frontline of fighting COVID-19, their perception of facility preparedness is particularly critical. Semann et al (2020) conducted a global cross-sectional study earlier on in the pandemic (between 24 March and 10 April 2020) documented the experiences of frontline RMNH workers in 81 countries and had 714 respondents (47% from LMICs including 16% from sub-Saharan Africa-SSA). The key results were that LMIC based respondents were worried about lack of access to evidence on COVID-19 in pregnancy, low perceived knowledge to care for COVID-19 maternity patients, a low proportion of institutions providing relevant COVID-19 training, and low availability of MNH COVID-19 guidelines. Additionally, almost 4 in 10 respondents reported substantially higher stress levels and there was a significant concern about MNH staff safety.^10^ Although this study had a relatively small sample size and did not explore the determinants of facility preparedness for RMNH services, it provided a snapshot of the preparedness, the response of various health systems, and the effect of COVID-19 pandemic in the initial phase of this evolving outbreak from health care providers perspective.

Since the Semann et al (2020) study, the WHO has produced guidelines for managing COVID-19 and for maintaining essential health services and these have been adopted by many countries in sub-Saharan Africa or similar ones produced to strengthen the responsiveness of their health system.^3^ The guidelines contains key recommendations to optimise health force capacity, including recruitment, repurposing within the limits of training and skills, redistributing roles among health workers while keeping health workers safe and providing mental and psychosocial support. It is unclear to what extent these guidelines have been implemented in health facilities and how this has affected health provider perception of their facility readiness to manage COVID-19. Other studies assessing preparedness to manage COVID-19 assessed health workers knowledge, attitude and practice, but did not explore the complex interaction between fear, anxiety, stress, support systems and health facility preparedness.^11–13^

The objective of this study was to assess the preparedness of the health system in Lagos State, Nigeria for the COVID- 19 outbreak based on the perception of RMNH providers, and to determine what factors (work-related stress, training, support and coping strategies/support mechanisms, availability of personal protective equipment (PPE) and guidelines, attendance for RMNH services) significantly predict this. It is expected that this study will generate context-specific data to improve the responsiveness of the health system for RMNH services and to minimise predicted adverse impact of the pandemic.

## Methods

A cross-sectional anonymous online survey of health workers providing RMNH services in Lagos State, Nigeria was conducted from 1^st^ to 21^st^ July 2020.

### Study site

Lagos State is the commercial nerve centre of Nigeria, located in the South-western region with an estimated population of over 20 million mostly urban residents and an annual growth rate of 3.2%. ^14^ The total fertility rate in the state is between 3.4 and 4.2 (national average 5.3), 86.4% of women used a skilled health provider for antenatal care ANC (national average 67%) and between 61 and 77% of women have health facility birth (national average 39%).^8^

Health is provided via health facilities owned by the Federal Government (Teaching hospitals and Federal Medical Centres), state government (state specialist, teaching and general hospitals), local government (primary health care facilities) and private/faith-based health care facilities. There are 306 primary healthcare centres, 27 registered general hospitals, 4, 421 private hospitals/specialist clinics/labs/diagnostic centres, and 5 tertiary health facilities. Most of these facilities provide RMNH services. ^15^

### Data collection tool

A pretested self-administered online questionnaire with six sections and 33 questions was used. Section 1 of the survey tool used contained study information and consent, and a question to confirm the provision of care in RMNH since March 2020. Section 2 collected demographic data such as gender, age group, professional cadre, type of facility and information on a management role if any. Section 3 collected information on the impact of COVID-19 on the availability and provision of RMNH services since March 2020, and reasons for these. Two of the sections had sub-scales for work-related stress and work-related burnout. Section 4 had six COVID-19 work-related stress questions, with four questions on a sub-scale with options 1 (not at all) to 5 (always/to a very high degree). Section 5 had seven COVID 19 work-related burnout questions, on a sub-scale with options 1 (never/almost never/to very low degree) to 5 (always/to a very high degree). The burnout questions were from the Copenhagen Burnout Inventory (CBI), a validated questionnaire with three sub-dimensions: Personal burnout, work-related burnout, and client-related burnout. The three separate parts of the CBI questionnaire were designed to be applied in different domains.^16^ Section 6 had eight questions on support and stress coping systems/mechanisms and overall perception of health facility readiness with options from 0 (unprepared to manage COVID-19 cases) to 5 (extremely well prepared to manage COVID-19 cases). In all, the data collection tool used in this study had 24 independent variables (17 categorical and 7 ordinal).

As part of the questionnaire development process, we analysed the reliability and internal consistency of both scales in the questionnaire using Cronbach alpha coefficient and the interitem correlation. Both scales have less than 10 questions, so a Cronbach alpha coefficient of 0.5 and inter-item correlation of 0.2-0.4 was acceptable. Any item in each subscale that adversely affected the Cronbach alpha coefficient was not included in the ordinal regression analysis.

### Data collection

A link to the survey was shared with health workers working in Lagos State via multiple social media platforms including Facebook and WhatsApp groups of health facilities and professional associations. Reminders were sent on these platforms every three days to maximise the reach of the survey.

### Data analysis

Four of the included independent variables were recoded so that the reference category or ordinal scale was appropriate: Availability of training on stress coping mechanism (unsure=0, No=1, Yes=2), concern about the availability of PPE and COVID-19 guidelines (extremely concerned=1, Unconcerned=5), worry about community discrimination/ stigma (extremely worried=1, Not worried=5), RMNH services unavailable at any time since March 2020 (Unsure=0, No=1, Yes=2).

We performed a descriptive analysis of independent variables: gender, age, professional cadre, facility type and management role and summarised outputs by frequency table, and cross-tabulation (facility type by the availability of RMNH services since March 2020, work colleague tested positive to COVID-19, off work due to suspected or confirmed COVID-19, work colleague COVID-19 related death, had COVID-19 training, availability of COVID-19 guidelines, availability of protocols for staff exposed to COVID-19 case and awareness of COVID-19 priority testing in a health facility). We used charts to describe the reasons for unavailability of, and reduced attendance for RMNH services between March and July 2020, as well as the frequency of coping/support mechanisms at health facilities. We also described the concern of health care workers about being infected at work, infecting family and friends, community stigmatisation/discrimination, availability of PPE and feeling of burnout.

We performed an ordinal regression analysis with health care worker perception of COVID-19 health facility preparedness (1-not prepared to 5-extremely well prepared) as the outcome or dependent variable and independent or predictor variables. We used the SPSS PLUM logistic regression and general linear models (GLM) programs for the ordinal regression analysis (IBM Corp. Released 2017. IBM SPSS Statistics for Windows, Version 25.0. Armonk, NY: IBM Corp). The assumptions of ordinal regression are the absence of multicollinearity and proportional odds were assessed. We assessed multicollinearity by calculating variance inflation factors (VIF). VIF values greater than 5 are of concern and 10 will suggest the presence of multicollinearity.^17^ We only included variables with VIF below 5 in the final logistic regression analysis.

The proportional odds assumptions, also known as the assumption of parallel lines, assess if the slope of the log-odds is equal for all categories of the dependent variable. If proportional odds cannot be assumed, then each predictor will have as many coefficients as thresholds in the ordinal regression. If the assumption of parallel lines is met, then only one coefficient needs to be calculated for each predictor.^18^

We used the ‘Model Fitting Information’ analysis to determine if the model improves our ability to predict the outcome/independent variable by comparing the -2 Log-likelihood of the Final model with the Intercept-only model. A statistically significant (p<0.05) chi-square statistic indicates that the Final model gives a significant improvement over the baseline intercept-only model. The overall model significance for the ordinal logistic regression was examined using the χ^2^ omnibus test of model coefficients (GLM analysis). McFadden’s *R*^2^ was examined to assess the per cent of variance accounted for by the independent variables (PLUM analysis) where values greater than 0.2 are indicative of models with excellent fit.^19^ Predicted probabilities of an event occurring were determined by Exp(B), also known as the odds ratios (GLM analysis).

### Ethical considerations and approval

We provided study information and frequently asked questions to all participants. Participants were required to consent to the study before completing the survey, they were free to withdraw their consent at any time. All incomplete surveys were excluded from the analysis. Ethics approval for the study was obtained from the Health Research and Ethics committee of College of Medicine University of Lagos (NHREC/19/08/2019B) and the Research and Ethics Committee of the Liverpool School of Tropical Medicine (20/052).

## Results

Three hundred and sixty-three health workers in Lagos state responded to the invitation to participate in the study, but 70.5% (256) were eligible to participate because they provided RMNH services since March 2020. All eligible participants consented to and completed the survey.

### Reliability of sub-scales in the questionnaire

Both sub-scales in the questionnaire were found to be reliable and internally consistent. The stress scale had 4 items, the Cronbach’s alpha was 0.743, the inter-item correlation 0.425, and the corrected item-total correlation for each item was above 0.4. The work-burnout scale had 7 items, we reverse coded one negative item on the scale (B04: energy for family and friends), the Cronbach’s alpha was 0.869, the inter-item correlation 0.470, and the corrected item-total correlation for each item was above 0.038.

### Descriptive statistics

Most respondents were female (72% or 184), aged 41-50 years (38% or 96), medical officers/registrars/house officers (40% or 101), from state Ministry of Health secondary facilities (52% or 133) and 63.7% (163) had a management role. **(Table 1)**

**Table 3:** Demographic characteristics of respondents (n=256)

| Variable | Categories | Frequency N (%) |
| --- | --- | --- |
| Gender | Female | 184 (72.4%) |
|  | Male | 70 (27.3%) |
|  | Not stated | 2 (0.8%) |
| Age (years) | 21-30 | 35 (13.7%) |
|  | 31-40 | 86 (33.6%) |
|  | 41-50 | 96 (37.5%) |
|  | 51-60 | 36 (14.1%) |
|  | Over 60 | 3 (1.2%) |
| Professional cadre | Community health officer/CRH worker | 23 (9%) |
|  | Consultant | 41 (16%) |
|  | Nurse/midwife | 89 (34.8%) |
|  | Medical officer/registrar/house officer | 101 (39.5%) |
|  | Others | 2 (0.8%) |
| Facility type | Primary care (Dispensary/PHC) | 39 (15.2%) |
|  | State Govt. secondary care (State hospital) | 133 (52%) |
|  | Fed. Govt. Tertiary care (FMC/Teaching hospital) | 60 (23.4%) |
|  | Private/Mission hospital | 23 (9%) |
| Management role | Yes | 163 (63.7%) |
|  | No | 93 (36.3%) |
| Management role type | HoD/dHoD | 45 (17.6%) |
|  | Medical Director/Chief Medical Officer/Medical Officer of Health/Chief Medical Officer | 28 (10.9%) |
|  | LG/State MoH rep. or health manager | 3 (1.2%) |
|  | CNO/Apex nurse. CHO/ward matron/hospital matron/coordinator/ward manager | 69 (27%) |
|  | Others | 18 (7%) |
|  | Not applicable | 93 (36.3%) |

### Availability of RMNH services between March and July 2020

Only 35.2% of the sample (90 respondents) reported that RMNH services were unavailable at any time between March and July 2020 in their HF. According to the HCPs sampled in our study, the two most common predominant contribution to reduced availability of RMNH services were community fear of infection from health facilities (37.8% or 34 respondents) and movement restrictions (30% or 27 respondents). Reduced availability of drugs and supplies and diversion of resources to treat COVID-19 patients were considered by the HCPs to have no effect on the availability of RMNH services **(Figure 1)**.

**Figure 1:**
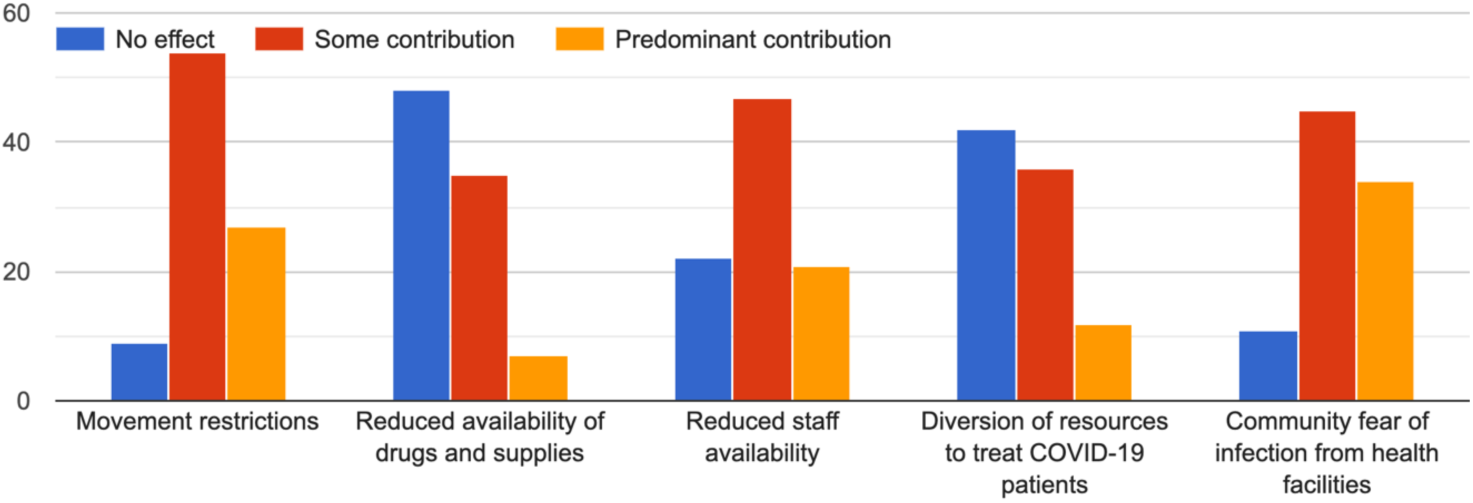
Reasons for unavailability of RMNH services since March 2020.

**Perceived effect of the COVID-19 outbreak on attendance for RMNH services if they were available since March 2020** Of the 125 respondents who reported that RMNH services were available, 97.6% (122) reported a reduction in attendance for these services (51.2% or 64) reported less than 50% reduction and 31.2% (39) reported more than 50% reduction in attendance).

The perceived contributions to reduced attendance for RMNH care where these services were available were similar to the reasons for non-availability of these services reported above. The two most common predominant contributions to reduced availability of RMNH services were community fear of infection from health facilities (47.6% or 49) and movement restrictions (47.6% or 49). Reduced availability of drugs and supplies and diversion of resources to treat COVID-19 patients were not considered by the HCPs to have affected the availability of RMNH services **(Figure 2)**.

**Figure 2:**
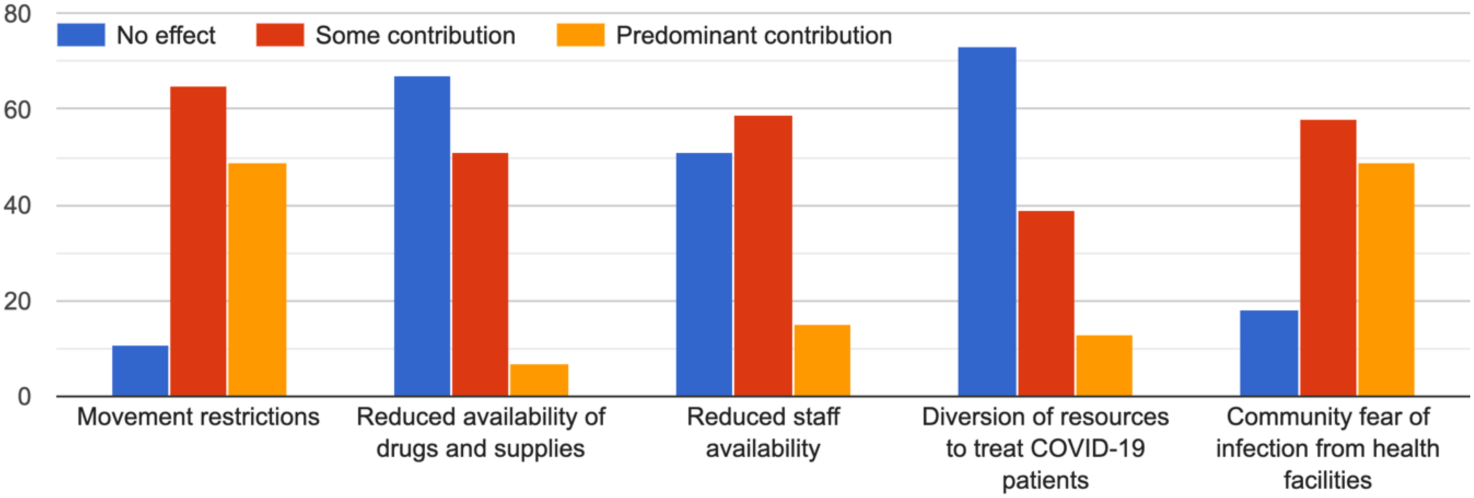
Reasons for reduced attendance for RMNH services.

### RMNH care provider perception of health facility readiness

About 88% (226) of RMNH care providers did not feel their health facility was sufficiently prepared. Eighty-seven per cent (117) and 91.7% (55) of all respondents from state secondary health facilities and federal government tertiary health facilities respectively reported that their facilities were not sufficiently prepared to manage COVID-19 cases. **(Table 2)**

**Table 2:** Variance Inflation Factors for independent variables included.

**Table 4:** Cross-tabulation of selected variables.

| Variable |  | Facility type (N (%)) |  |  |  |  |  |
| --- | --- | --- | --- | --- | --- | --- | --- |
|  |  | PHC | State MoH<br>secondary care<br>hospital | FMOH/SMoH<br>tertiary care<br>hospital | Private/Faith<br>-based/NGO<br>hospital | Others<br>(Research<br>Institute) | Total |
| RH services unavailable since March 2020 | No | 26 (66.7%) | 73 (54.9%) | 13 (21.7%) | 12 (52.2%) | 1 (100%) | 125 (48.8%) |
|  | Yes | 8 (20.5%) | 37 (27.8%) | 37 (61.7%) | 8 (34.8%) | 0 (0.0%) | 90 (35.2%) |
|  | Unsure | 5 (12.8%) | 23 (17.3%) | 10 (16.6%) | 3 (13%) | 0 (0.0%) | 41 (16%) |
| Off-work due to suspected or confirmed C-19 infection | No | 31 (79.5%) | 96 (72.7%) | 39 (65%) | 18 (78.3%) | 0 (0.0%) | 185 (72.5%) |
|  | Yes | 8 (20.5%) | 36 (27.3%) | 21 (35%) | 5 (21.7%) | 1 (100%) | 70 (27.5%) |
| Work colleague positive to COVID-19 | No | 24 (61.5%) | 43 (32.3%) | 13 (21.7%) | 14 (60.9%) | 0 (0.0%) | 94 (36.7%) |
|  | Yes | 15 (38.5%) | 90 (67.7%) | 47 (78.3%) | 9 (39.1%) | 1 (100%) | 162 (63.3%) |
| Colleague off-work due to suspected or confirmed C-19 infection | No | 20 (51.3%) | 41 (30.8%) | 11 (18.3%) | 5 (21.7%) | 0 (0.0%) | 77 (30.1%) |
|  | Yes | 19 (48.7%) | 92 (69.2%) | 49 (81.7%) | 18 (78.3%) | 1 (100%) | 179 (69.9%) |
| Work colleague C-19 related death | No | 36 (92.3%) | 118 (88.7%) | 54 (90.4%) | 22 (95.7%) | 1 (100%) | 231 (90.2%) |
|  | Yes | 3 (7.7%) | 15 (11.3%) | 6 (10%) | 1 (0.4%) | 0 (0.0%) | 25 (9.8%) |
| Attended C-19 training | No | 9 (23.1%) | 36 (27.1%) | 14 (23.3%) | 8 (34.8%) | 0 (0.0%) | 67 (26.2%) |
|  | Yes | 30 (76.9%) | 97 (72.9%) | 46 (76.7%) | 15 (65.2%) | 1 (100%) | 189 (73.8%) |
| Availability of C-19 guidelines | No | 12 (32.8%) | 21 (30.8%) | 7 (11.7%) | 5 (21.7%) | 1 (100%) | 45 (17.6%) |
|  | Yes | 27 (69.2%) | 27 (69.2%) | 53 (88.3%) | 18 (78.3%) | 0 (0.0%) | 211 (82.4%) |
| Availability of protocols for staff exposed to positive C-19 case | No | 16 (41%) | 46 (34.6%) | 14 (23.3%) | 9 (39.1%) | 0 (0.0%) | 85 (33.2%) |
|  | Yes | 23 (59%) | 87 (65.4%) | 46 (76.7%) | 14 (60.9%) | 1 (100%) | 171 (66.8%) |
| Awareness of C-19 priority testing in HF | No | 17 (43.6%) | 74 (55.6%) | 23 (38.3%) | 13 (56.5%) | 1 (100%) | 127 (49.6%) |
|  | Yes | 22 (56.4%) | 59 (44.4%) | 37 (61.7%) | 10 (43.5%) | 0 (0.0%) | 129 (50.4%) |
| Overall HF COVID-19 preparedness | Insufficiently prepared | 87.2% (34) | 88% (117) | 91.7% (55) | 82.6% (19) | 1 (100%) | 226 (88.3%) |
|  | Sufficiently prepared | 12.8% (5) | 12% (16) | 8.3% (5) | 17.4% (4) | 0 (0.0%) | 30 (11.7%) |

### Concern about being infected at work, infecting family and friends, community stigmatisation/discrimination, availability of PPE and feeling of burnout

Forty per cent (104) of respondents were moderately or extremely worried about community stigmatisation or discrimination as a result of their potential exposure to COVID-19 cases in their facilities and 39% felt moderate or extreme work-related burnout, since the COVID-19 outbreak. Sixty-six per cent (158), 70.4% (180) and 84% (205) of respondents were moderately or extremely concerned about being infected at work, exposing family members and friends to COVID-19, and availability of PPE and related guidelines at work respectively.

### Support systems to cope with providing care during the pandemic

Most of the respondents (74.2% or 190) had received training on how to respond to COVID-19 and 83.2% (213) had guidelines for providing RMNH services during the pandemic 67.6% (173) but only 49.6% (127) were aware of any priority COVID-19 testing for health workers (**Table 2**).

About 54% (137) of respondents reported empathy from HF management, 46.8% (120) had been trained on coping with stress and improving mental health since the outbreak, 46.4% (119) reported the availability of counselling services and 43.4% (111) reported that resources to cope with stress and improve mental health such as mobile apps, documents and websites since the outbreak had been shared at their facility **(Figure 3)**.

**Figure 3:**
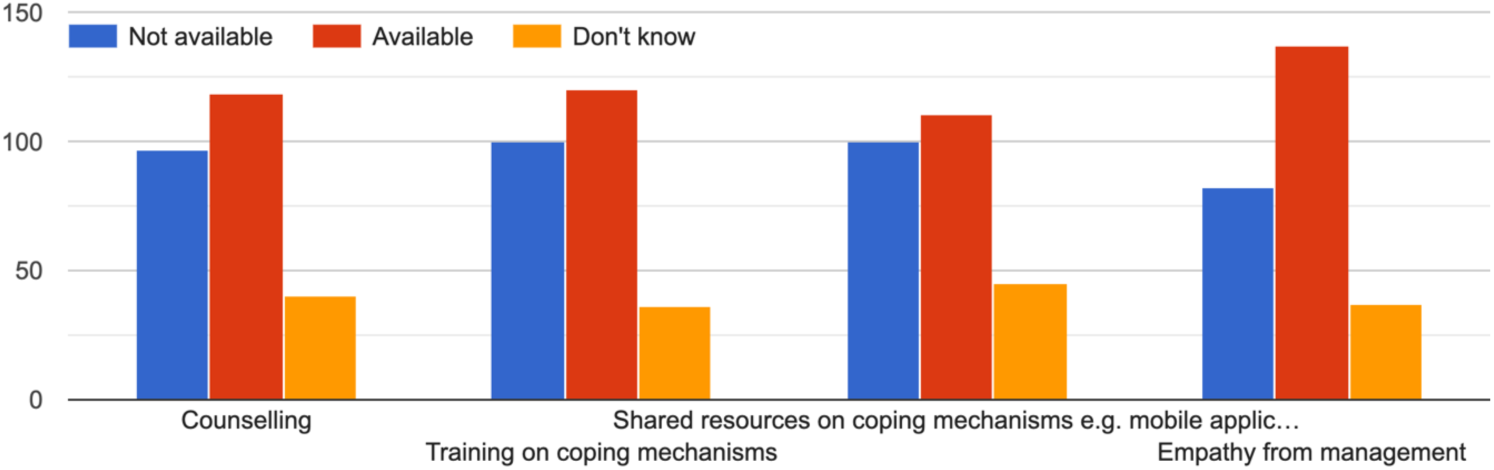
Frequency of coping/support mechanisms available at health facilities.

Other coping mechanisms for work-related stress included prayer 69.1% (177), discussion with colleagues and family members 71.5% (183), rest and meditation 66% (169), exercise 30.5% (78) and engaging with support groups via social media 75 (29.3%).

### Ordinal Logistic Regression

An Ordinal Logistic Regression was conducted to determine if the odds of observing each response category of health worker perceived HF preparedness could be explained by the variation in the independent variables included (**Table 1**).

## Compliance with ordinal regression analysis assumptions

### Multicollinearity

Of the 24 independent variables, thirteen independent variables (3 ordinal and 10 nominal) included in the regression analysis have VIFs less than 5, we conclude that there is no high correlation between the independent variables included in the ordinal regression analysis.

### Test of parallel lines

The null hypothesis states that the location parameters or slope coefficients are the same across response categories, therefore lines of the same slope are parallel. The -2 loglikelihood of the null hypothesis is 576.102, for the general model 530.554, X^2^ 45.548, DF=72, p=0.994. Therefore, we failed to reject the null hypothesis and the proportional odds assumption of our model holds.

### Model fitting

The -2 Log-likelihood for a model with intercept only (a model that does not control for any predictor or independent variable and simply fits an intercept to predict the outcome or dependent variable) was 764.685 while that for the Final model was 576.102, X^2^ of 188.583, DF=24 p<0.001. Therefore, we reject the null hypothesis and conclude that the final model gives a significant improvement over the baseline intercept-only model. This was consistent with the results of the Ominus test from the GLM analysis: the full model was a significant improvement in fit over the null hypothesis (no predictors) [X^2^ (24) = 188.583, P<0.001]

### Pseudo R-square (Link function-Logit)

The Link function-Logit McFadden’s R square and Nagelkerke value were used as measures of ‘Goodness of Fit’ because the outcome variable output was normally distributed. McFadden R-squared value calculated for this model was 0.242. Nagelkerke value was 0.547, meaning that up to 54.7% of the variation in the outcome variable is explained by the model.

### Parameter estimates

The test of model effects (GLM analysis) showed that only 2 independent variables had a statistically significant effect on the level of perception of facility preparedness, likelihood ratio Chi-square (*p*-value): concern about the availability of PPE and COVID-19 guidelines 15.430, df: 4 (*p*=0.004), and the level of satisfaction with HF management communication on COVID-19 87.941, df: 4 (p<0.001) (

An increase in the health worker concern about the availability of PPE and COVID-19 guidelines was associated with an increase in odds of satisfaction with the level of the health care facility preparedness. The regression coefficient for any level of satisfaction less than extremely satisfied with HF management communication was significant, *B* = -1.932— 5.806, χ^2^ = 19.339-61.155, *p* <0.001, suggesting that a one-unit increase in the level of satisfaction would increase the odds of observing a higher category of health facility COVID-19 preparedness with an odds ratio of 0.79-2.92 (**Table 5)**.

**Table 5:** Variance Inflation Factors for independent variables included.

| **Variable** | **VIF** |
| --- | --- |
| Off work due to suspected or confirmed COVID-19 | 1.37 |
| Work colleague tested positive for COVID-19 | 2.62 |
| Concern about the availability of PPE and guidelines | 1.55 |
| Availability of protocols for staff exposed to COVID-19 case | 1.69 |
| Level of satisfaction with HF communication on COVID-19 | 2.60 |
| Availability of COVID-19 HF priority testing for health workers | 1.34 |
| Attended COVID 19 training | 1.24 |
| Work colleague died from COVID 19 disease | 1.19 |
| Colleague off work due to COVID 19 | 2.39 |
| RMNH services unavailable at any time since March 2020 | 1.28 |
| Worries about stigma/discrimination related to COVID-19 | 1.85 |
| Availability of RMNH COVID 19 guidelines | 1.43 |
| Availability of training on stress coping mechanisms | 1.80 |

**Table 6:** Tests of Model Effects.

| Tests of Model Effects |  |  |  |
| --- | --- | --- | --- |
| Source | Type III |  |  |
|  | Likelihood Ratio Chi-Square | df | Sig. |
| *Availability of training on stress coping mechanisms | 1.041 | 2 | 0.594 |
| Worries about stigma/discrimination related to COVID-19* | 1.553 | 4 | 0.817 |
| Off work due to suspected or confirmed COVID-19 | 0.437 | 1 | 0.509 |
| Colleague off work due to COVID 19 | 1.449 | 1 | 0.229 |
| Work colleague tested positive C19 | 0 | 1 | 0.986 |
| Work colleague died from COVID-19 | 0.131 | 1 | 0.717 |
| Attended COVID 19 training | 0.018 | 1 | 0.892 |
| Availability of RMNH COVID 19 guidelines | 3.819 | 1 | 0.051 |
| Availability of protocols for staff exposed to COVID-19 case | 0 | 1 | 0.995 |
| Availability of COVID-19 HF priority testing for health workers | 3.664 | 1 | 0.056 |
| *Availability of PPE and COVID-19 guidelines | 15.43 | 4 | **0.004** |
| Level of satisfaction with HF communication on COVID-19 | 87.941 | 4 | **0** |
| *RMNH services unavailable at any time since March 2020 | 5.542 | 2 | 0.063 |
| **Dependent Variable: Level of satisfaction with health facility preparedness** |  |  |  |
| Model: (Threshold), Availability of training on stress coping mechanisms, Worries about stigma/discrimination related to COVID-19, Off work due to suspected or confirmed COVID-19, Colleague off work due to COVID 19, Availability of RMNH COVID 19 guidelines, Availability of protocols for staff exposed to COVID-19 case, Availability of COVID-19 HF priority testing for health workers, Availability of COVID-19 HF priority testing for health workers, Availability of RMNH COVID 19 guidelines, Level of satisfaction with HF communication on COVID-19, Level of satisfaction with HF communication on COVID-19 |  |  |  |
| *recoded variables |  |  |  |

**Table 7:** Ordinal Logistic Regression Results for independent variables predicting HCW perception of COVID-19 health facility preparedness.

|  | B | Std Error | 95% Wald Confidence Interval |  | Hypothesis testing |  |  | Exp(B) | 95% Wald Confidence Interval for Ex (B) |  |
| --- | --- | --- | --- | --- | --- | --- | --- | --- | --- | --- |
|  |  |  | Lower | Upper | Wald Chi-Square | df | Sig. |  | Lower | Upper |
| Threshold |  |  |  |  |  |  |  |  |  |  |
| Satisfaction with HF preparedness = Extremely unsatisfied | -6.335 | 1.0214 | -8.337 | -4.333 | 38.468 | 1 | 0 | 0 | 0 | 0.013 |
| Satisfaction with HF preparedness = Very unsatisfied | -4.07 | 0.9897 | -6.01 | -2.131 | 16.916 | 1 | 0 | 0.02 | 0.002 | 0.119 |
| Satisfaction with HF preparedness =Neither satisfied or unsatisfied | -1.935 | 0.9658 | -3.828 | -0.042 | 4.013 | 1 | 0.045 | 0.14 | 0.022 | 0.959 |
| Satisfaction with HF preparedness =Very satisfied | 0.383 | 0.9433 | -1.466 | 2.232 | 0.165 | 1 | 0.685 | 1.47 | 0.231 | 9.32 |
| Availability of training on stress coping mechanisms = Unsure | 0.167 | 0.3997 | -0.616 | 0.951 | 0.175 | 1 | 0.675 | 1.18 | 0.54 | 2.588 |
| *Availability of training on stress coping mechanisms=No | -0.208 | 0.3187 | -0.832 | 0.417 | 0.424 | 1 | 0.515 | 0.81 | 0.435 | 1.517 |
| *Availability of training on stress coping mechanisms=Yes | 0<sup>a</sup> | . | . | . | . | . | . | 1 | . | . |
| Worries about stigma/discrimination related to COVID-19=extremely worried | 0.22 | 0.4114 | -0.587 | 1.026 | 0.285 | 1 | 0.593 | 1.25 | 0.556 | 2.79 |
| Worries about stigma/discrimination related to COVID-19 =very worried | 0.568 | 0.4628 | -0.339 | 1.475 | 1.506 | 1 | 0.22 | 1.77 | 0.712 | 4.372 |
| Worries about stigma/discrimination related to COVID-19=Neither worried or unworried | 0.276 | 0.4003 | -0.509 | 1.06 | 0.474 | 1 | 0.491 | 1.32 | 0.601 | 2.887 |
| Worries about stigma/discrimination related to COVID-19=Some worry | 0.26 | 0.4156 | -0.555 | 1.074 | 0.391 | 1 | 0.532 | 1.3 | 0.574 | 2.928 |
| Worries about stigma/discrimination related to COVID-19=Not worried | 0<sup>a</sup> | . | . | . | . | . | . | 1 | . | . |
| Off work due to suspected or confirmed COVID-19=No | -0.205 | 0.3099 | -0.812 | 0.403 | 0.436 | 1 | 0.509 | 0.82 | 0.444 | 1.496 |
| Off work due to suspected or confirmed COVID-19=Yes | 0<sup>a</sup> | . | . | . | . | . | . | 1 | . | . |
| Colleague off work due to COVID 19=No | 0.491 | 0.4088 | -0.31 | 1.292 | 1.442 | 1 | 0.23 | 1.63 | 0.733 | 3.641 |
| Colleague off work due to COVID 19=Yes | 0<sup>a</sup> | . | . | . | . | . | . | 1 | . | . |
| Work colleague tested positive C19=No | 0.007 | 0.4093 | -0.795 | 0.81 | 0 | 1 | 0.986 | 1.01 | 0.452 | 2.247 |
| Work colleague tested positive C19=Yes | 0<sup>a</sup> | . | . | . | . | . | . | 1 | . | . |
|  |  |  | Lower | Upper | Wald Chi-Square | df | Sig. |  | Lower | Upper |
| Work colleague died from COVID-19=No | 0.156 | 0.4314 | -0.689 | 1.002 | 0.131 | 1 | 0.717 | 1.17 | 0.502 | 2.723 |
| Work colleague died from COVID-19=Yes | 0<sup>a</sup> | . | . | . | . | . | . | 1 | . | . |
| Attended COVID 19 training=Yes | 0.04 | 0.2928 | -0.534 | 0.614 | 0.018 | 1 | 0.892 | 1.04 | 0.586 | 1.847 |
| Attended COVID 19 training=No | 0<sup>a</sup> | . | . | . | . | . | . | 1 | . | . |
| Availability of RMNH COVID 19 guidelines =No | -0.746 | 0.3829 | -1.496 | 0.004 | 3.796 | 1 | 0.051 | 0.47 | 0.224 | 1.004 |
| Availability of RMNH COVID 19 guidelines =Yes | 0<sup>a</sup> | . | . | . | . | . | . | 1 | . | . |
| Protocols for exposed staff available=No | -0.002 | 0.3201 | -0.629 | 0.626 | 0 | 1 | 0.995 | 1 | 0.533 | 1.869 |
| Protocols for exposed staff available=Yes | 0<sup>a</sup> | . | . | . | . | . | . | 1 | . | . |
| HF priority C-19 testing available=No | -0.53 | 0.2777 | -1.074 | 0.014 | 3.642 | 1 | 0.056 | 0.59 | 0.342 | 1.014 |
| HF priority C-19 testing available=Yes | 0<sup>a</sup> | . | . | . | . | . | . | 1 | . | . |
| *Extremely concerned about PPE and guideline Availability=1 | -0.232 | 0.8457 | -1.89 | 1.425 | 0.075 | 1 | 0.784 | 0.79 | 0.151 | 4.159 |
| *Very concerned about PPE availability and Guidelines=2 | 0.92 | 0.8798 | -0.805 | 2.644 | 1.092 | 1 | 0.296 | 2.51 | 0.447 | 14.067 |
| *Neither concerned nor unconcerned about PPE and guideline availability=3 | 0.522 | 0.9022 | -1.246 | 2.29 | 0.335 | 1 | 0.563 | 1.69 | 0.288 | 9.879 |
| *Some concern about PPE and guidelines availability=4.00 | 1.072 | 0.9672 | -0.824 | 2.968 | 1.229 | 1 | 0.268 | 2.92 | 0.439 | 19.45 |
| *Unconcerned about PPE and guidelines availability=5.00] | 0<sup>a</sup> | . | . | . | . | . | . | 1 | . | . |
| Extremely unsatisfied with HF management Communication=5 | -5.806 | 0.7425 | -7.262 | -4.351 | 61.155 | 1 | 0 | 0 | 0.001 | 0.013 |
| very unsatisfied with HF management communication=4 | -4.125 | 0.5718 | -5.245 | -3.004 | 52.038 | 1 | 0 | 0.02 | 0.005 | 0.05 |
| Neither satisfied or unsatisfied with HF management communications=3 | -3.591 | 0.4831 | -4.537 | -2.644 | 55.244 | 1 | 0 | 0.03 | 0.011 | 0.071 |
| Very satisfied with HF management communication=5 | -1.932 | 0.4394 | -2.793 | -1.071 | 19.339 | 1 | 0 | 0.15 | 0.061 | 0.343 |
| Extremely satisfied with HF management communication=5 | 0<sup>a</sup> | . | . | . | . | . | . | 1 | . | . |
| *RMNH services unavailable = Unsure | -0.84 | 0.3878 | -1.601 | -0.08 | 4.697 | 1 | 0.03 | 0.431 | 0.202 | 0.923 |
|  |  |  | Lower | Upper | Wald Chi-Square | df | Sig. |  | Lower | Upper |
| *RMNH services unavailable =Yes | -0.043 | 0.2754 | -0.583 | 0.497 | 0.025 | 1 | 0.875 | 0.958 | 0.558 | 1.643 |
| *RMNH services unavailable = No | 0<sup>a</sup> | . | . | . | . | . | . | 1 | . | . |
| (Scale) | 1<sup>b</sup> |  |  |  |  |  |  |  |  |  |
| **Dependent Variable: Level of satisfaction with health facility preparedness** |  |  |  |  |  |  |  |  |  |  |
| Model: (Threshold), Availability of training on stress coping mechanisms, Worries about stigma/discrimination related to COVID-19*, Off work due to suspected or confirmed COVID-19, Colleague off work due to COVID 19, Availability of RMNH COVID 19 guidelines, Availability of protocols for staff exposed to COVID-19 case, Availability of COVID-19 HF priority testing for health workers, Availability of COVID-19 HF priority testing for health workers, Availability of COVID-19 HF priority testing for health workers, Availability of RMNH COVID 19 guidelines, Level of satisfaction with HF communication on COVID-19, Level of satisfaction with HF communication on COVID-19*<br>*recoded variables<br>Set to zero because this parameter is redundant.<sup>a</sup><br>Fixed at the displayed value.<sup>b</sup> |  |  |  |  |  |  |  |  |  |  |

The regression coefficient for any level of concern about the availability of PPE and COVID-19 guidelines less than unconcerned about the availability of PPE and COVID-19 guidelines was significant, *B* = 0.92-1.07, χ^2^ = 19.339-61.155, *p* <0.001, suggesting that a one-unit increase in the level of concern about the availability of PPE and COVID-19 guidelines would increase the odds of observing a higher category of satisfaction with health facility COVID-19 preparedness an odds ratio of 0. 02-0.15. (**Table 5**)

## Discussion

### Summary of main findings

Of the two hundred and fifty-six RMNH service providers respondents, most worked in public-owned health facilities (91%), were female (72%), nurse/midwives (35%), non-specialist medical doctors (40%) and had both management and clinical roles (64%). About 35% of respondents reported that RMNH services were unavailable at some point between March 2020 and July 2020, mostly in the tertiary public hospitals. Less than a third of RMNH workers were off work due to suspected or confirmed COVID-19 infection and this was a lot more in secondary and tertiary public-owned hospitals, compared to primary health care and faith-based/private owned health facilities. Most of the respondents who work in secondary and tertiary hospitals reported that a colleague had tested positive for COVID-19 compared to those who work in primary care or faith-based/NGO hospitals. Almost three-quarters of respondents reported that COVID-19 training was available at their health facilities, this was more available in tertiary public owned hospitals. Sixty-three per cent, 70.4% and 84% of respondents were moderately or extremely concerned about being infected at work, exposing family members and friends to COVID-19, and availability of PPE and related guidelines at work. About 88% (226) of RMNH care providers did not feel their health facility was sufficiently prepared.

Our final model was a statistically significant predictor of RMNH worker perception of facility preparedness after controlling for the independent variables, explaining up to 54.7% of the variation in the outcome variable. A one-unit increase in the level of satisfaction with the communication from HF management and the level of concern about the availability of PPE and COVID-19 guidelines would increase the odds of observing a higher category of satisfaction with HF COVID-19 preparedness (OR 0.79-2.92, p<0.001 and 0.02-0.15 p<0.001 respectively).

### Interpretation

We set out to investigate the determinants of perceived health facility preparedness, using ordinal regression analysis controlling for known risk factors for work-related stress, burnout, personal preparedness (COVID-19 training) and institutional preparedness (availability of relevant guidelines, PPE, staff support systems).

About 88% (226) of RMNH health workers did not feel their health facility was sufficiently prepared, the availability of PPE and related guidelines and satisfactory communication from HF management were significant determinants of this. Similar results were reported from one study earlier in the pandemic. In a cross-sectional study conducted in Jordan conducted in March 2020 that included only frontline medical doctors treating COVID-19 cases, doctors having institutional protocols for dealing with COVID 19 cases and those with sustained availability of PPE reported higher scores of self preparedness to manage COVID-19 cases.^20^ Following the publication of WHO guidelines to support the maintenance of essential health services in the context of COVID-19, we expected different results 4 months into the pandemic.

Health workers in Lagos expressed serious concerns about being infected at work or infecting family members/friends and this is similar to studies reported during previous infectious disease outbreaks and earlier on during this outbreak.^10,21,22^ Such expressed fears and concerns typically lead to stress. High levels of health worker work-related stress have been reported in systematic reviews before and during the COVID-19 pandemic. ^7,23^ The risk factors for COVID-19 related physical and mental health impacts include working in a high-risk department, diagnosed family member, improper PPE use. Most RMNH care providers are female and Shaukat et al. (2020) also reported that female health workers and nurses were disproportionately affected by the mental health impacts of COVID-19.^7^ Female health workers may have additional burden during this pandemic, caring for their families, providing care without adequate protection and support. Before the COVID-19 pandemic, Onigbogi and Banerjee (2019) reported work overload, poor communication and lack of resources and equipment were significant risk factors for psychological stress in Nigerian health workers.^23^

Preparedness can be viewed at two levels, individual preparedness and institutional preparedness. But the institution typically drives preparedness, for example making training available to staff is likely to increase individual preparedness, providing relevant protocols, protective supplies such as PPE and support mechanisms for health workers to improve individual resilience, prevent burnout, stress and actively support staff who suffer stress and psychological effects associated with providing care during an infectious disease outbreak. Compared to earlier on in the pandemic, training has improved but shortages PPE and lack of guidelines for its use are persisting problems.^10,12,13,20^ The improvement in training is expected following the publication of the WHO operational guidance for maintaining essential health services for the COVID-19 context and the push from WHO, Africa CDC, Nigeria CDC and the state and Federal Ministries of Health for staff training. However, this can be improved, so that all health workers are regularly trained, this is critical for self-confidence, preparedness and resilience and should be complemented by adequate supplies of PPE and support systems.

Our results may suggest that the WHO guidelines for maintaining essential health services have not been fully implemented or is not having an impact on the perceived preparedness of the HFs to manage COVID-19 cases and maintain essential health services. The link between, concern for PPE availability and communication from HF management regarding COVID-19 guidelines was evident from the ordinal regression analysis, and underscores the importance of caring for the carers, during a large disease outbreak. Generally, the knowledge of Nigeria health workers on COVID-19 has increased over time, but they need to be continually briefed and informed, covering not only up to date clinical management but also covering recognition, management and support strategies for work-related stress and burnout.^24^ The consequences of not taking action are huge, with health worker stress, burnout, and psychological problems associated with COVID-19, being some reported in our study. This is a wake-up call for health system planners.

Individual-led stress coping method/strategies reported by respondents included, prayer (69.1%), meditation (66%), peer support (29.3%) and exercise (30.5%) are not uncommon in low resource countries. Witter et al (2017) reported the use of similar coping strategies by health workers in a post-conflict and infectious disease outbreak countries.^25^ While the room to implement such individual-led coping mechanisms must be provided, including praying/meditation rooms, the role of institutional support for frontline health workers should not be minimised. Institutional-led interventions such as psychological first aid to all health workers, adequate workforce planning, regular debriefing, stress and burnout surveillance, access to psychologists and counsellors, clear and consistent communication have been recommended to minimise the risk of stress and burnout during such outbreaks.^21,26,27^ Some of these interventions were reported as available by over half of the respondents, there may be room for improvement to proactively plan and consistently implement them in all health facilities during such outbreaks.

Our finding that RMNH services were unavailable at some point since public health and social measures were implemented in Lagos State is consistent with a nationally representative telephone survey conducted in April 2020, by the Partnership for Evidence-Based Response to COVID-19 (PERC) that a high proportion of respondents who need health care have had difficulties accessing such services (39%). The same study found that adherence to staying at home order (implemented from 29^th^ March 2020 in Lagos State) completely adhered to by only 23% of the population, while it is clear that movement restrictions will have affected utilisation of essential health services, health care worker anxiety about the risk of infection, community stigmatisation may have affected the provision of services as well. Community fear of infection from health facilities was the second most common reason given by health workers for none utilization or low attendance for RMNH services. In the subsequent PERC survey reported in September 2020, nearly three-quarters of respondents felt confident that they will get medical help if they fell ill. The change in public perception as the pandemic progressed may be associated with increasing knowledge, attitude and practice, and lower COVID-19 risk perception by the public.^24, 28^ Our finding of reduced attendance for RMNH services has been reported in the earlier multicounty cross-sectional study, however, with increased public knowledge, and reduced risk perception attendance for RMNH services should be expected and health facilities should be prepared for this.^10^ It was interesting to note that in our study, diversion of resources to treat COVID-19 patients and reduced availability of medicines and supplies were perceived as having no effect or significant contribution to non-utilisation or reduce attendance for RMNH services. This is contrary to the global expectations at the start of the outbreak, especially with the shutdown of international transport systems that is key for the importation of medicines and supplies.^3,29^ Nigeria largely depends on imports of pharmaceuticals from India and China, but there may have been sufficient stock levels of essential medicines at the initial phase of the outbreak.^29^ A more plausible explanation may be that as utilization reduced, demand for medicines and medical supplies also reduced, as such RMNH workers never appreciated the potential impact.

### Strengths and limitations

There are key strengths of note in our study, this was the first to go beyond just reporting levels of preparedness to understanding the determinants of health worker perception of HF preparedness and mechanisms that they use to cope during the COVID-19 pandemic. Compared to the earlier studies on preparedness to manage COVID-19 we explored the interaction between determinants of preparedness that included known risk factors of health worker stress and factors that can improve preparedness using ordinal regression analysis. Our study was conducted in a COVID- 19 safe way (online survey), most respondents were in management and clinical roles and were front line health workers, thus increasing the validity of the results.

However, our study is not without limitations, Our sample was relatively small (256) for the size of the health workforce but was more than the minimum sample size of 240 required for ordinal regression analysis based on the recommendation of Hosmer, Lemeshow, and Sturdivant (2013) of a minimum sample for ordinal regression analysis of 10 observations per independent variable. ^30^ Also faith-based/private owned health facilities were under-represented (less than 10% all response) but over 85% of health facilities in Lagos are privately owned. ^15^ Other studies have reported a preference for privately owned health facilities for childbirth by about 50% of women irrespective of social class. ^31,32^ Therefore, our study may not be representative of the preparedness of private health facilities from the perspective of health workers and a similar study with a large sample of private and faith-based facilities is needed to understand this.

## Conclusion

Training of RMNH care providers, provision of PPE, guidelines, provision of support and coping support systems complemented with appropriate communication from health facility management are likely to improve the capacity of HFs to provide quality care during the COVID-19 outbreak. Similar studies are needed to evaluate the perception of preparedness in private health facilities, including the perspective of women and their carers, this is important to improve overall health system preparedness during an outbreak and for quality improvement. Full implementation of the WHO operational guidance for maintaining essential health services for the COVID-19 context should be prioritised, including monitoring of recommendations for optimising the health workforce for a satisfactory level of health facility preparedness during this pandemic.

## Data Availability

The datasets used and/or analysed during the current study are available from the corresponding author on reasonable request.

## Availability of data and materials

The dataset used and/or analysed during the current study are available from the corresponding author on reasonable request.

## Authors’ contributions

ABT, BM, MCC, ABB: study design, interpretation and review. CA: conceptualisation, study design, analysis, interpretation, drafting and review.

## Acknowledgements

We are immensely grateful to Dr Adesola Pitan (Deputy Director of Medical Services, Lagos State Health Service Commission), Dr Kehinde Okunade (Secretary, Society of Gynaecology and Obstetrics of Nigeria, Lagos sector), Dr Japhet Olugbogi (Chairman, Committee on Covid19 Nigerian Medical Association, Lagos), Dr Anthonia Onyenwenyi (Deputy Director, CHO Training School, LUTH), Comrade Blessing Israel (Chairman National Association of Nigeria Nurses and Midwives Lagos State Council), Dr Doyin Ogunyemi (Secretary Association of Public Health Physician of Nigeria, Lagos Chapter), and Dr Ibironke Sodeinde (Acting President, Medical Women’s Association of Nigeria, Lagos Chapter) for their immense support in disseminating the survey tool used in this study.

